# Prevalence and factors associated with iron deficiency anaemia among preterm infants attending clinics in Dodoma, Tanzania: A facility-based cross sectional study

**DOI:** 10.1101/2024.10.25.24316098

**Authors:** Christer R Mchwampaka, Dina C. Mahamba, Shubi Matovelo, Shakilu Jumanne

## Abstract

**Aim:** Iron deficiency anemia (IDA) among preterm infants is a global public health problem due to its effects on development, immunity and general growth that may be irreversible. Despite the high burden of preterm births in Tanzania the prevalence and factors associated with IDA among preterm infants remains relatively understudied and this constituted of the aim of this study.

**Methods:** A cross-sectional analytical study design was conducted among 190 preterm infants who attended pediatric clinic in Dodoma City from December 2022 to May 2023. Socio-demographic, clinical information, and laboratory markers of hemoglobin and iron status were obtained. Logistic regression analysis was applied to determine factors associated with the outcome (Iron Deficiency Anaemia).

**Results:** A total of 190 preterm infants were enrolled in the study and the mean age was 4.01 months (SD ±0.99 month. The prevalence of IDA among preterm infants was 11.58%. Factors associated with IDA were very low birth weight (AOR 6.906, CI: 1.4774-32.359, p *value <* 0.0142), preterm infant not supplemented with Iron (AOR 6.282, CI:1.045-37.763*, p value<* 0.0446, multiple pregnancies (AOR 6.848, CI:1.692-27.708, *p value <*0.0006) and severe anaemia during pregnancy (AOR 11.998, CI:5.068-40.197, *p value<* 0.0001).

**Conclusion:** Iron deficiency anemia was found to be 11.58% which fall under public health problem under WHO classification. To reduce IDA among preterm infants, there has to be an emphasis on iron supplementation to all preterm infants, and those with very low birth weight, born from mother who had multiple pregnancy and severe anaemia during pregnancy need close follow up and improved postnatal

**Keynotes:**

- This study enrolled 190 preterm infants attending pediatric clinic in Dodoma city to determine factors associated with iron deficiency anaemia (IDA)
- Globally IDA in preterm infants range between 25 to 80, which complicates their prematurity by causing poor growth, poor functioning of multiple organ systems, poor neurological development and contributes to death and disability
- Iron supplementation to all preterm infants is highly recommended

## Background

Iron deficiency anemia (IDA) develops when the body’s iron reserves are insufficient to maintain the regular synthesis of red blood cells (RBCs), inadequate dietary iron, impaired iron absorption, bleeding, or loss of body iron in the urine may be the cause (Hempel & Bollard, 2016) IDA among preterm infants is documented as a global public health problem, with prevalence ranging from 25% to 80%(Ferri et al., 2014)This is consistent with world health organization (WHO)’s standard, revealing that when the IDA prevalence is 5% it is considered a public health burden (Paulley & Duff, 2022). Preterm infants in the first week of life had 26.4 times likely to develop iron depletion which can lead to IDA compared to term infants with normal birth weights, the high prevalence of IDA among preterm infants has been associated with low iron stores at birth, early onset of erythropoiesis, rapid catch-up growth, iatrogenic blood loss, limited dietary sources of iron, (Moreno-Fernandez et al., 2019). IDA in preterm infants complicates their prematurity by causing poor growth, poor functioning of multiple organ systems, and poor neurological development (McCarthy et al., 2019a).This leads to the increase of infant mortality rates (Moreno-Fernandez et al., 2019)

In sub-Saharan Africa, IDA is not well reported especially for preterm infants, most of the studies have reported the prevalence of IDA for under five children accounting to 60% (Lemoine & Tounian, 2020),In East Africa according to a study done by Hellen in Kenya to assess the prevalence of iron deficiency and iron deficiency anaemia in low-birth-weight infants on follow -up at Kenyatta National Hospital was 14.5% (Hellen G, 2019). In Tanzania few conducted studies demonstrated (44.2%) prevalence of IDA among infants in Dar es Salaam (Omar Lweno et al., 2022) and 50% among under-five age children in Kilimanjaro (Urio et al. 2019).

The factors that were documented to be associated with IDA among preterm infants include Birth weight and phlebotomy (Beard et al., 2007; Ganjigunta et al., 2021; Strauss, 2010) Gestation type (Ru et al., 2016; Shinar et al., 2017), Age (Hassan et al., 2016) feeding practices preterm infants and mothers who are not supplemented with iron during pregnancy, Low hemoglobin level, maternal illness (Gurung et al., 2020b: (Q.Li et al.,2019)

In Tanzania, only few studies have been conducted and the factors were generalised to infants and under five year’s children not specific to preterm infants, the factors stated included; maternal hemoglobin, sex of the child, low income, low birth weight, small for gestational age (SGA), preterm delivery, and dietary status of infants and women (Omar Lweno et al., 2022). This study aimed to determine the prevalence and factors associated with iron deficiency anaemia in preterm infants attending premature clinic in Dodoma city.

## Methods

This was a hospital based analytical cross section study design conducted at pediatric clinic at Dodoma Region Referral Hospital (DRRH) and RCH clinic in Makole health centre in Dodoma city, central Tanzania. Duration of study was six months starting from December 2022 to May 2023.The average attendance of preterm per months at paediatric clinic is 88 at DRRH, and 32 per months at RCH clinic at Makole Health centre. Paediatric clinic at DRRH serves infants who have been discharged from the DDRH and therefore infants from other facilities before reaching the target weight of 2.5kg. At DRRH, each visit to the clinic, preterm infant’s weight and vital signs are carefully monitored to track their progress. Additionally, the pediatric clinic ensures that the preterm infants receive appropriate supplementation by providing iron and multivitamin supplements,blood investigations like full blood picture, blood grouping and cross matching are taken from infants whose assessment findings by attending doctor dictate. RCH clinic at Makole Health Centre receive large population of infants’ average of 40 for immunization according to their age as per Expanded Programme on Immunization.

### 3.7 The sample size

The sample size was calculated using the Kish & Leslie

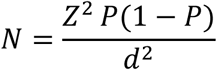

Whereby:

`o` N = sample size
`o` Z = score for 95% Confidence Interval which is 1.96
`o` P = prevalence in a previous study
`o` d = tolerable error set at 5%,

P = Prevalence of IDA in preterm infants with low birth weight from previous study which was 14.5% was used according to study done in Kenya at Kenyatta National Hospital in 2019 (Hellen G, 2019)

The study included preterm infants aged 3 to 6 months, (infants born before < 37 completed weeks) Confirmed by RCH card 4

Infants with feeding difficulties due to different medical conditions example congenital anomalies like cleft lip and palate, hydrocephalus, history of bleeding, who were sick were, (eg high temperature ≥38C) were excluded

Preterm infants meeting the inclusion criteria were consecutively enrolled until the required sample size was attained

Enrolment was done only after parents/guardians had signed a written informed consent at admission in the neonatal ward. Ethical approval to conduct the study was granted by the University of Dodoma research and ethical review board with ethical clearance reference number MA.84/261/59/154. Permission to conduct the study was sought from the DRRH and Makole Health centre administration. A pretested structured data collection sheet was used to extract demographic, medical history, clinical data of preterm infants and mothers, enrolled in the study. Physical examination and laboratory results was done on all preterm infants up component of the study. Social demographic data such as place of residence, age, sex, gestation age, weight, current feeding practices, phlebotomy and iron supplement status were recorded, also parents’ or guardians’ Gestation type, last hemoglobin level before delivery, any maternal illness like pregnancy induced hypertension, gestation diabetics and postpartum hemorrhage were collected. Each participant was examined for paleness and any sign of iron deficiency anemia like brittle or spoon-shaped nails.

Under aseptic technique peripheral vein was identified and tourniquet applied 4-5 inches from the puncture site, by using a gauge needle, the skin was punctured to access the vein. Approximately 6 milliliters of venous blood drawn. 4 milliliter was kept in the red top blood Collection tube for CRP and serum ferritin test, 2milliliter were kept on an EDTA purple top blood collection tube for Full Blood Picture test. All samples were kept in ice bag 2-8 centigrade, then transported to the lab according to laboratory instructions. (FBP) was measured using CELL-DYN Ruby hematology (Genway Biotech, USA) whereas serum ferritin and CRP were measured using Snibe Maglumi 800 analyser (Genway Biotech, USA). From the FBP; Hb level. MCV, and MCH were determined. All the samples were tested at DRRH laboratory, samples collected from Makole Health Center were collected and transported to DRRH Laboratory through Cool box temperature of 2C to 8C. IDA was defined if they met any of the following criteria: HB < 10, MCV < 80fl, MCHC <32 g/ dl, (Serum Ferritin < 12 ug/L and CRP < 5 mg/dL)

Data analysis was done using the Statistical Package for Social Science (SPSS) software version 25. Categorical variables were summarized using percentages (%) and frequency distributions, whereas mean and median with their measures of distributions were used to summarize continuous variables. Multinomial logistic regression was used to determine the association between clinical and laboratory factors and adverse outcomes for IDA. Variables with a p-value of < 0.2 for unadjusted multinomial logistic analysis were included in the adjusted multinomial logistic regression analysis to determine independent factors associated with IDA poor outcomes. Odds ratios were reported with 95% confidence interval and variables with p values of <0.05 were considered statistically significant.

### Results

From December 2022 to May 2023, A total of 190 enrolled in this study 139 from DRRH and 51 from Makole Health Centre

**Figure 1:**
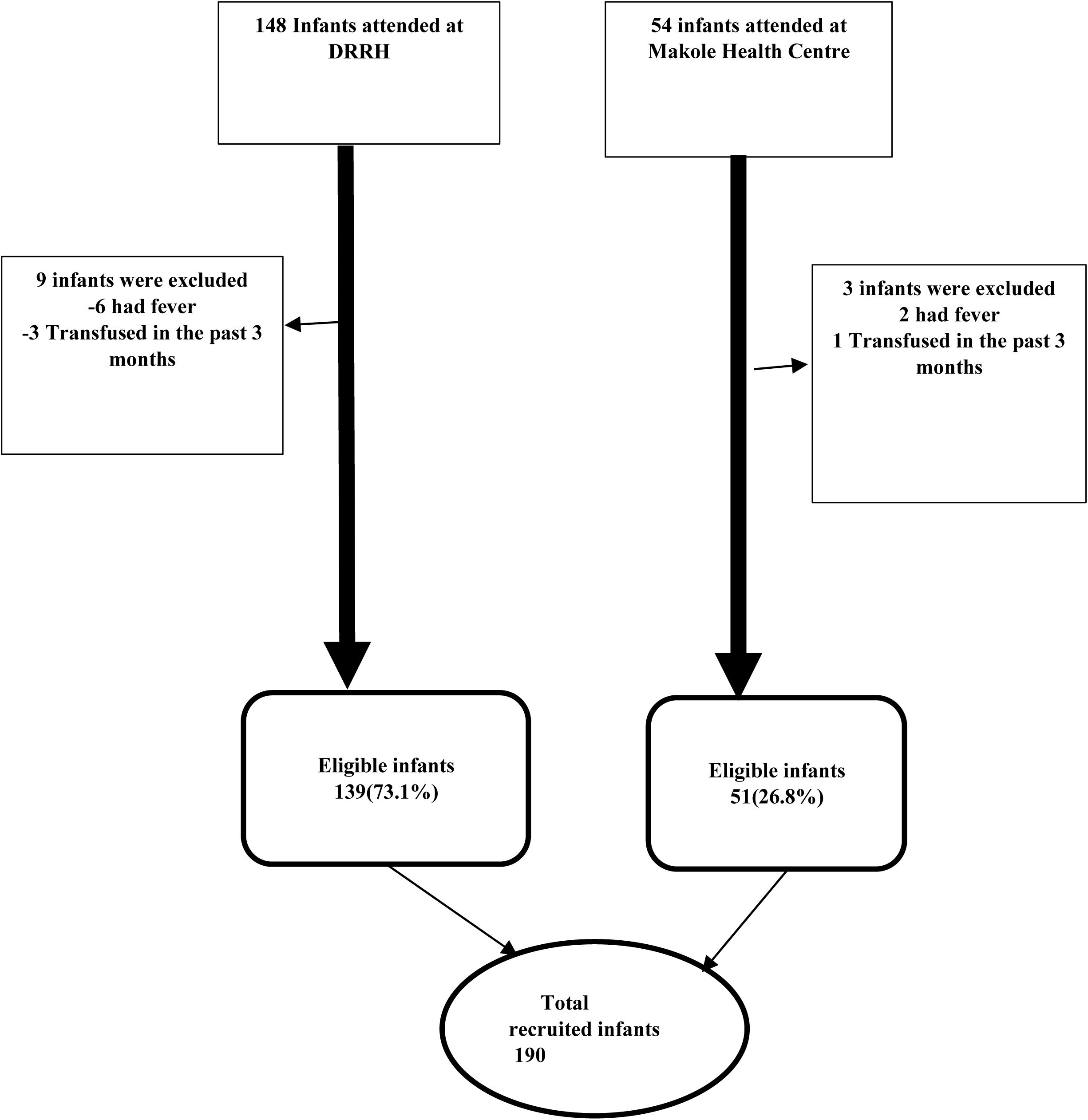
Flow chart of Enrolled in preterm infants.

## RESULTS

A total of 190 preterm infant with response rate of were enrolled in this study. 139 from pediatric clinic at DRRH and 51 from Makole Health were enrolled in this study

### Infant and maternal demographic data and clinical characteristics

Majority of preterm infants 109, (57.37%) were females. Mean age was 4.01 months (SD 0.99 month). Majority of infants (157, (82.63%) were born as singletons, more than half 123 (64.74%) had birth weight of 1.5 kg or more and 175 infants (92.11%) were on iron supplements. Most 145(90.65%) of the mothers had no maternal illness (pregnancy induced hypertension, gestation diabetics and postpartum hemorrhage) during pregnancy and 22(11.58) had severe anaemia. **Table 1** summarizes the demographic data characteristics of the infants and maternal

**Table 1:**
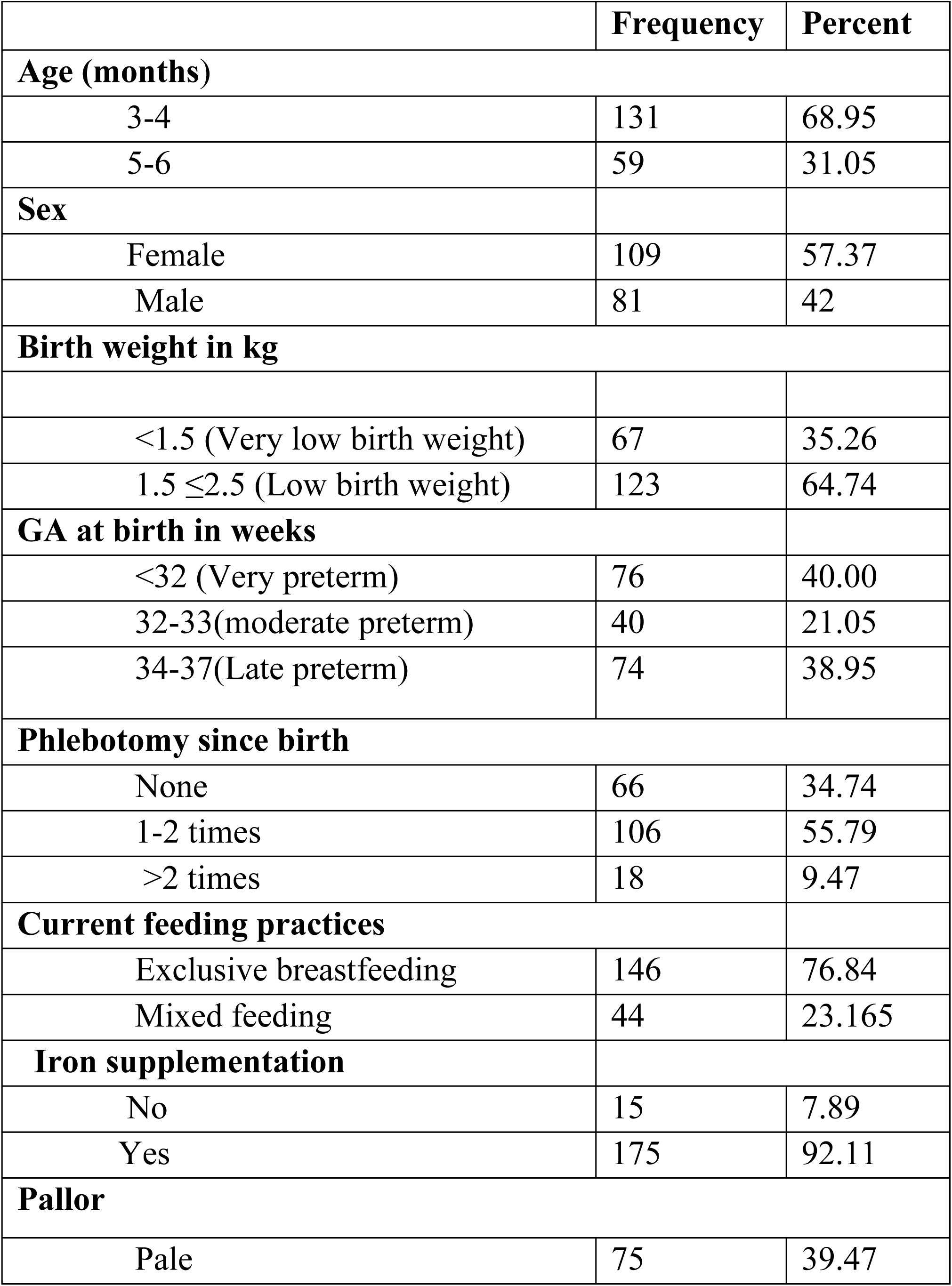
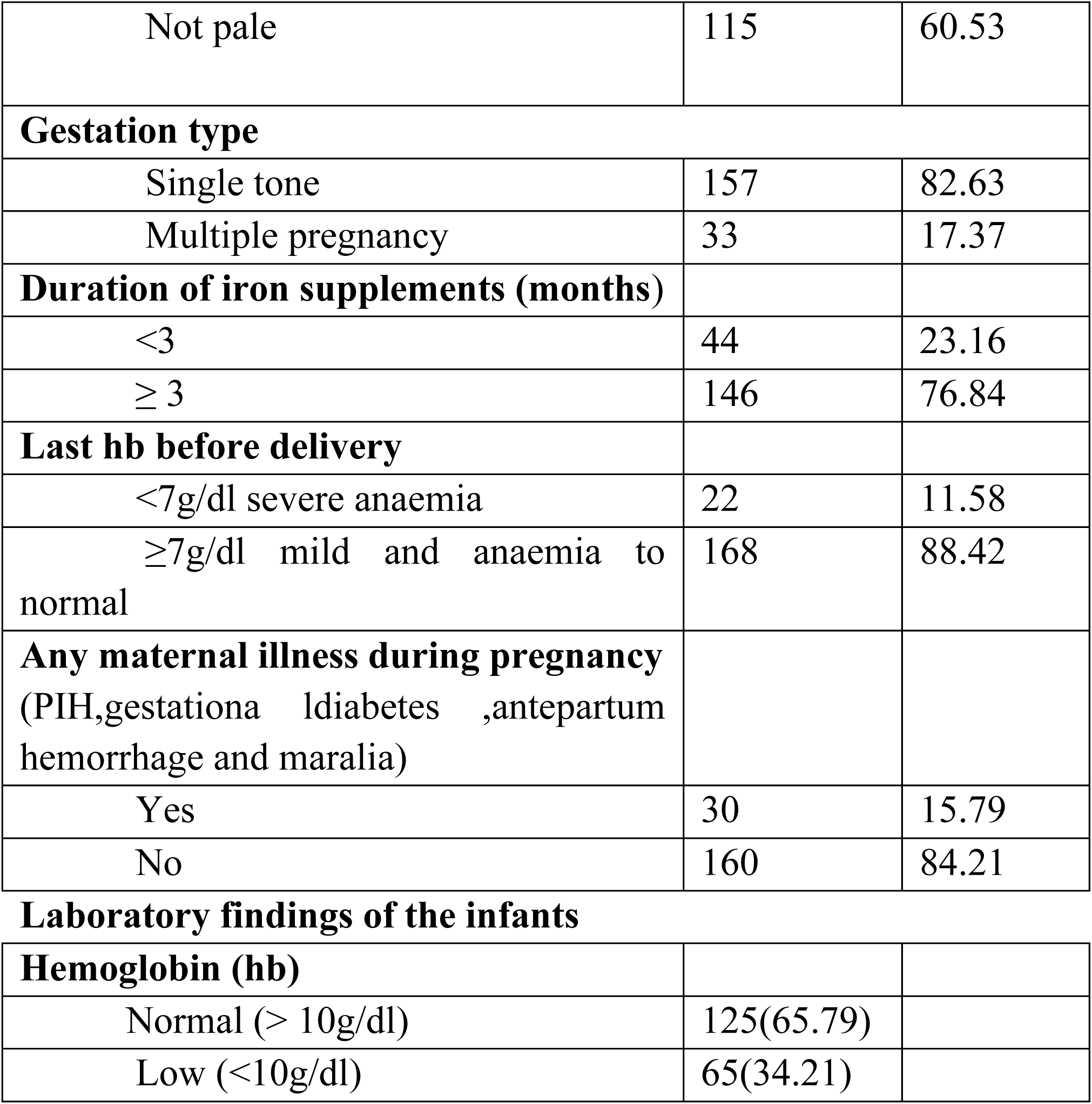
Infant and maternal demographic and clinical characteristics by IDA.

### Prevalence of IDA among infants born preterm

The total of twenty-two of the 190 study participants had IDA making overall prevalence of IDA among preterm unfants to be 11.58%

**Figure 1:**
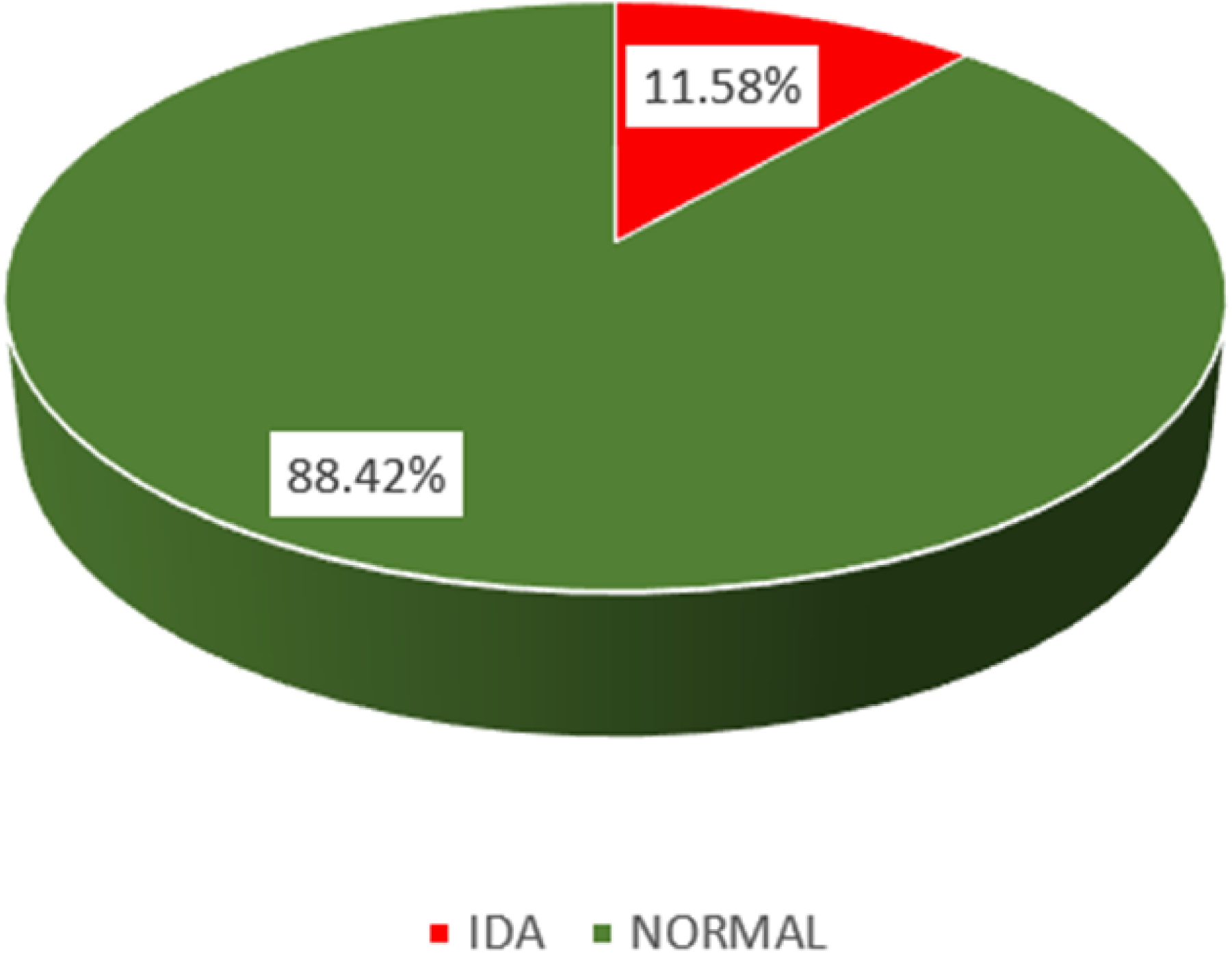
Prevalence of Iron deficiency anaemia among preterm infants.

### Factors associateds with Iron deficiency anemia among preterm infants aged 3 to 6 months

Chi-square test was used to assess the factors associated with iron deficiency anemia (IDA) among preterm infants (3 to 6 months). In this study the findings show, that IDA was associated with very low birth weight (p=<0.0001), preterm infants who are not on iron supplementation (p=. <.0001), very preterm (gestation age at birth of less than 32 weeks) (p= 0.0165), multiple pregnancy (p=0.007), duration of Iron supplementation of maternal of less than 3 month (p=0.0084) and severe anemia during pregnancy (p=0.0011). See table 2 for further observation of the findings

**Table 2:**
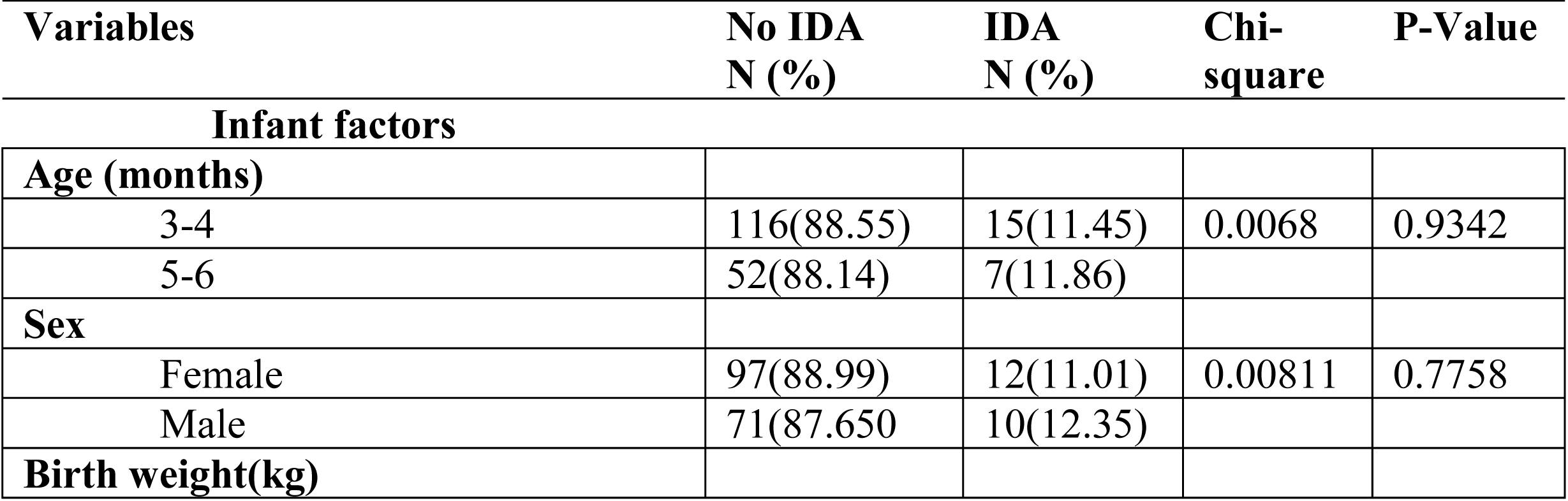
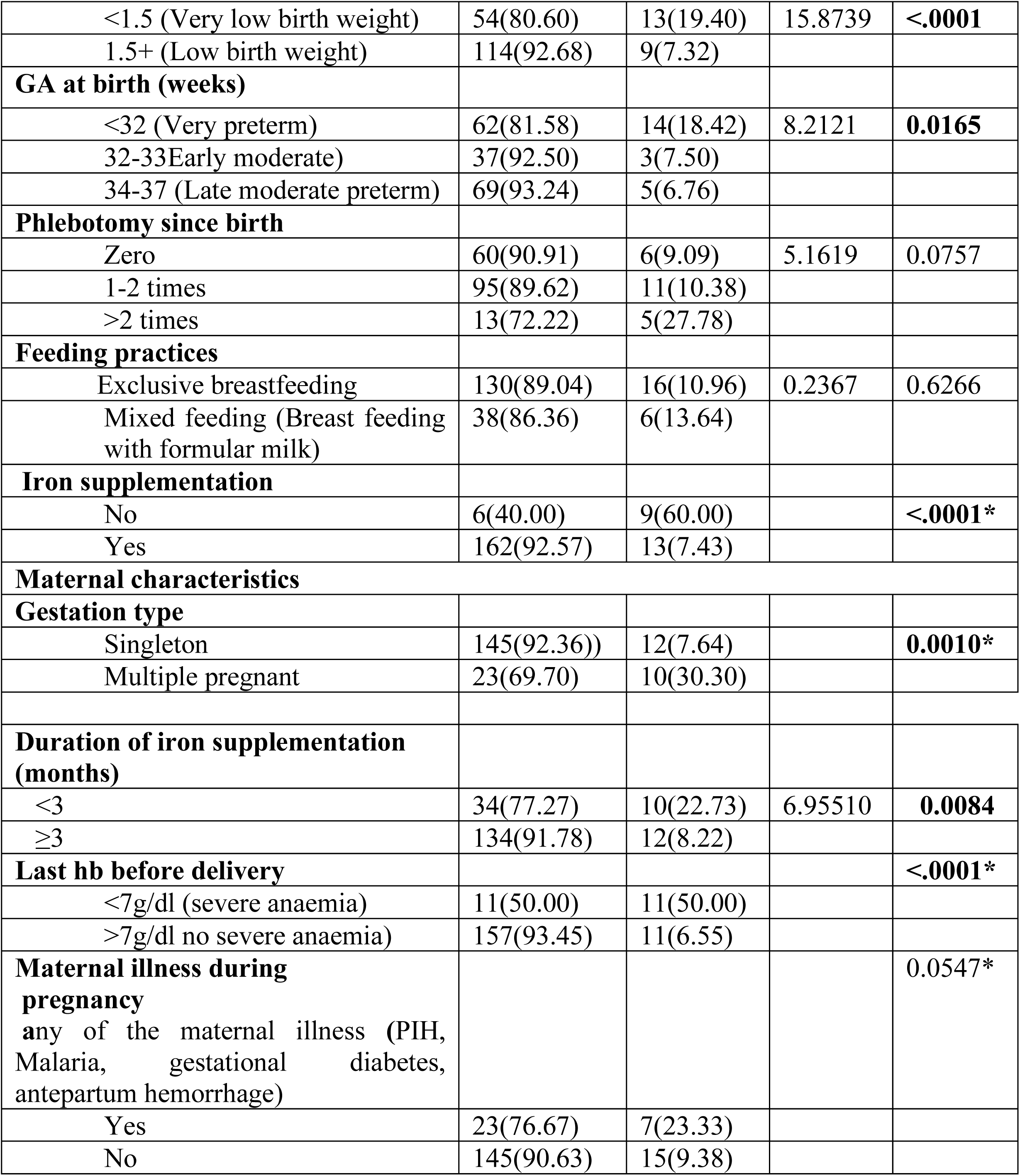
Chi-square test and results on the assessment of factors associated with iron deficiency anemia (IDA) among preterm infants (aged 3 to 6 months)

### 4.5 Factors associated with iron deficiency anemia and preterm infants aged 3 to 6 months

Binary logistic regression was used to determine the factors association with iron deficiency anemia among preterm infants aged 3 to 6 months. which revealed that; Very low birth weight (OR6.80, *p value <0.0003),* Low gestation age (OR3.14, *p value <0.0.028),* having more phlebotomy (OR3.85, *p value <0.047),* Preterm Infant not supplemented with Iron (OR18.69, *p value <0.000),* mothers who had multiple pregnancy (OR5.26, *p value <0.001),* Less than 3 months on iron supplemented with Iron during pregnancy (OR3.29, *p value <0.011),* Low Hb for mothers before delivery (OR14.27, *p value <0.000),* maternal illness during pregnancy (OR2.94, *p value <0.034).* All those factors with p value <0.25 at bivariate level were subjected to multivariate model.

Upon adjusted binary logistic regression; only four factors with p value <0.05 at 95% CI remained significant associated with IDA, the factors included: Very low birth weight (OR6.91, *p value <0.014),* preterm infant not supplemented with Iron (OR6.28, *p value <0.045),* mothers who had multiple pregnancy (OR6.85, *p value <0.007)* and Low Hb for mothers before delivery (OR11.99, *p value <0.001)*.

**Table 3 summarizes the factors associated with iron deficiency anaemia**.

**Table 3:**
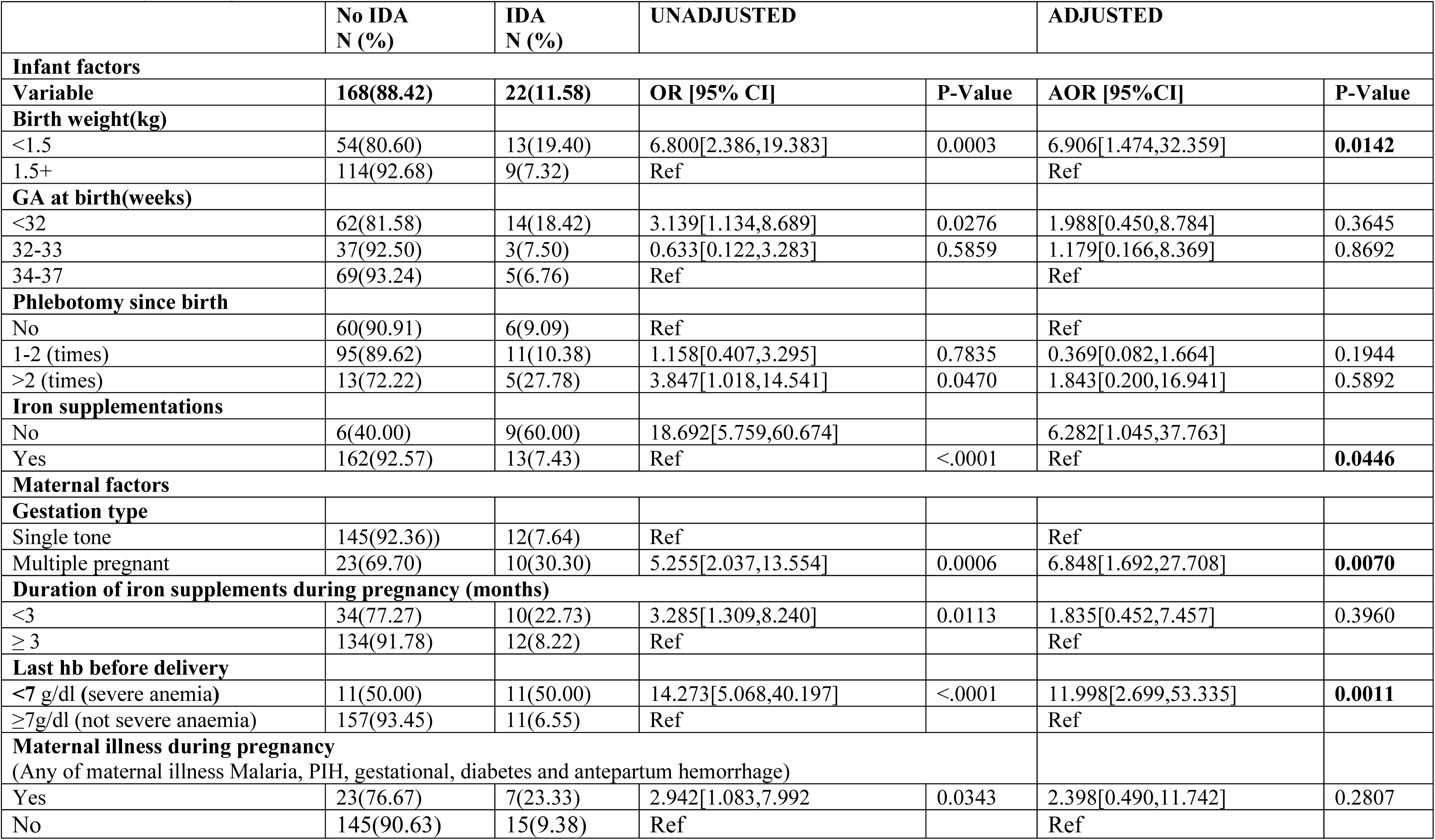
Logistic Regression for Factors Associated with Iron Deficiency Anaemia for Preterm Infants Aged 3 to 6 Months.

## CHAPTER FIVE DISCUSSION

Iron deficiency anemia (IDA) leads to poor growth, poor functioning of multiple organ systems, poor neurological development, effects on memory, cognition and auditory brain responses and irreversible long-term complications such as bone diseases among preterm infants. In Tanzania few conducted studies demonstrated high (44.2%) prevalence of IDA among infants in Dar es Salaam (Omar Lweno et al., 2022).Prevention is achieved by early initiation of iron supplementation of elemental iron for preterm infants as prophylaxis against IDA. In this cross-section study 190 preterm infants were enrolled. The study aimed to determine the prevalence and factors associated with IDA in preterm infants aged 3 to 6 months. The prevalence of IDA was found to be 11.58%. Very low birth weight, (birth weight less than 1.5kg, those who were not supplemented by iron, and those mothers who had multiple gestation and severe anemia were strongly associated with occurrence of IDA.

The overall prevalence of iron deficiency anemia (IDA) in this study was found to be 11.58%. This is a public health problem as define by WHO that if prevalence ≥5% (measuring by ferritine concentrations below the recommended cut off values (Paulley & Duff, 2022). However it is almost similar to the study that was conducted in Sweden at Umea University that found the prevalence of IDA to be 9.9%(Berglund et al., 2010). Similarity in findings may be attributed by age groups of preterm infants in both studies that were within 6 months following birth, all infants had low birth weight and majority in both groups were exclusive breastfeeding. The findings in this study were also close to another study done in Kenya that revealed prevalence of 14.6% for IDA (Hellen W Githaiga, 2019). Similarity in the prevalence could be possibly due to the fact that both studies involved children with low birth weight, same study design, majority of infants were on iron supplements, similar culture setting, availability of health services and majority of infants were 6 months and below same as current study.

The prevalence of this study was low compared to the study conducted in Turkey which found the prevalence of Iron deficiency anemia in late-preterm infants to be 42.8% (Ozdemir et al., 2013). This observed differences in findings can be attributed by the fact that participants involved in Turkey were not on iron supplement unlike this study where by majority of participants (92.57%) were on iron supplementation, this might have led to the higher prevalence in the study conducted Turkey since the documented evidince indicate that premature infants who are not supplemented are associate with IDA (Ozdemir et al., 2010). In addition, the study in Turkey included, lower age premature infants (2 to 4 months) that utilize and need more Iron for their growth compared to 3 to 6 months age used in our study. Postnatal, iron stores can be rapidly depleted during the first six to eight weeks, coinciding with the onset of erythropoiesis and rapid catch-up growth (Rao & Georgieff, 2009). Additionally, another study conducted in Brazil reported a higher prevalence of 26.5% among preterm infants with very low birth weight aged one year of corrected age (Ferri et al., 2014).The difference may be attributed by difference in the birth weight for the study population where the study focused on infants with less than 1.5 kg and gestation age of <34 weeks compared to the current study that considered infants with less than 2.5 kg and gestation age <37 weeks. The documented evidence indicate that higher demand of Iron in infants with low birth weight is due to rapid growth, also most of the iron transfer to fetus do occurs during the third trimester, therefore the level of IDA increase as gestation age decrease, (Jopling et al., 2014)

Furthermore the finding in the current study was higher compared to the study conducted in Indonesia which revealed the prevalence of 6% (Puspitasari et al., 2017). This difference probably is due to involvement of participants born at very preterm (gestation age of 28 weeks to the current study compared that of Indonesia, which involve moderate to rate preterm (gestation age of 32 <37) as documented that there is associated between IDA and low gestation age this is due to increase of rapid growth,high iron needs and reduced iron store(Omar Lweno et al., 2022).

Basing on the findings of the current study, preterm infants with very low birth weight (less than 1.5kg) were six-fold more likely to have IDA compared to their counterparts (above 1.5kg) This result is comparable to others studies conducted in Korea, India and Brazil where their results indicated a positive association of low birth weight with IDA (Ganjigunta et al., 2021; Joo et al., 2016; Ferri et al.,2014). Different studies demonstrated a correlation between a lower birth weight and IDA in infants, this is mainly attributed by lower store of iron at birth and higher iron requirements due to increased postnatal growth (Moreno-Fernandez et al., 2019).Iron is an essential element for the function of growing and differentiating cells so if there is in rapid growth as seen in very low birth more iron is needed compare to low birth weight (Ferri et al., 2014)

In this study we observed that preterm infants who had iron supplementation were less likely to develop IDA. This result is comparable to other studies conducted in Indonesia and China which revealed similar correlation (Berglund et al., 2010; Li et al., 2021). Also a systematic study that evaluated iron supplementation in preterm and low-birth-weight infants, confirmed that iron supplementation increased hemoglobin and ferritin concentrations and a reduction in iron deficiency anemia (McCarthy et al., 2019b). Another study conducted in Italy that found similar correlation went further to explain the association basing on the fact that the more the duration of iron supplementation, the higher the possibility of increased hemoglobin and ferritin concentrations and a reduction in iron deficiency anemia, (Raffaeli et al., 2020). It has also been documented that maternal iron is transferred to fetus during the third trimester of gestation but once the transfer is interrupted by preterm birth iron stored deprived, thus preterm infants that are not supplemented with iron end up with IDA(Berglund et al., 2010).

The current study revealed that mothers who had low hemoglobin level during pregnancy increased the odds of having preterm infants with IDA. This finding is similar to the study done in Philippines, of which the results indicated that at 6 months evidence from the 2015–16 TDHS-MIS cross-sectional household survey in Tanzania (Msaki et al., 2022), which found a similar result and is supported by other evidence that suggested that anemic pregnant women are more likely to have preterm infant (Gurung et al., 2020), and hence increased the risk for IDA. In addition other research evidence conclude that preterm infants are deprived of the significant iron deposit that occurs in the third trimester of pregnancy and have reduced iron stores at birth compared with term infants (McCarthy et al., 2019b). This effect is more severe when the woman had anemia in pregnancy especially in the third trimester, the observed result and association could be linked to maternal anemia during pregnancy which has correlation to having infant with LBW(Enawgaw et al., 2019; Figueiredo et al., 2019). Another study conducted at Era Lucknow Medical College in India (Shukla et al., 2019) on effect of maternal anemia on the status of iron stores in infants, concluded that maternal IDA may have an effect on the iron stores of newborns. During pregnancy,the mother’s body prioritizes transferring iron to the developing fetus.As a result,mother with anemia may have lower iron stores herself,and this can affect the iron stores that are transferred to the baby in utero.(Terefe et al., 2015)

It was also uncovered by this study that mothers who had multiple pregnancy were more likely to have preterm infants with IDA compared to those who had a singleton pregnancy. The current study results concurs with what was reported in the study conducted at the Ohio State University Wexner Medical Center in USA which concluded that twin babies born preterm (≤37 weeks) are at greater risk of low iron stores at birth and of ID later in infancy (Campbell et al., 2022). It could further be linked to the fact that iron deficiency and anemia are prevalent in women with multiple pregnancy (Ru et al., 2016), so that for pre-term twin babies, the odds for IDA increases as found in our study. The maternal iron requirements are increased during twin pregnancies, estimated to be 1.8 times more than during singleton pregnancies, due to increased foetal and placental needs as well as increased maternal plasma volume expansion and red blood cell mass (Shinar et al., 2017). Therefore, compared to singleton gestations, maternal hemoglobin (Hgb) in multiple pregnancies is lower in all trimesters, with an estimated IDA rate of 2.4 to even 4 times (Shinar et al., 2017)

### 6.1 Conclusion

This study revealed that Iron Deficiency Anaemia is prevalent among preterm infants aged 3 to 6 months at health facilities in Dodoma city despite majority of them being on haematinics supplement. The prevalence was found to be 11.58%, the very low birth weight of the preterm infants, preterm infant not supplemented with Iron, mothers who had multiple pregnancy and mothers having low Hb before delivery were associate with IDA. Emphasis on iron supplementation to all preterm infants, and those with very low birth weight, born from mother who had multiple pregnancy and severe anaemia during pregnancy need close follow up and improved postnatal to reduce IDA.

## Data Availability

All relevant data are within the manuscript and its Supporting Information files.

## Acknowledgements

We cannot express enough gratitude to the women who consented to let their preterm infants to participate in the study. Special thanks to Dr Halima Kasimu in charge of pediatric clinic from DRRH and Sr Josephine Dikoko incharge from Makole health centre and all the staff at the at premature clinicat DRRH and RCH clinic.

## Conflict of interest

The authors report no conflict of interest

CRM: Conceptualization of the project, data collection and analysis and preparation of the manuscript draft

DM: Conceptualization of the project, data analysis and preparation of the final manuscript SM: Conceptualization of the project, data analysis and preparation of the final manuscript SJ: Conceptualization of the project, data analysis and preparation of the final manuscript

## Abbreviations

DRRH: Dodoma Regional Referral Hospital
EDTA: Ethylene Diaminetetraacetic Acid
FBP: Full Blood Picture
RCH: Reproductive and Child Health
DRRH: Dodoma Regional Referral Hospital
g/dl: Gram per Deciliters
Hb: Hemoglobin
VLBW: Very Low Birth Weight
WHO: World Health Organization

